# Influence of Workplace Happiness on Organizational Commitment among Healthcare Workers in Ibadan Metropolis, Oyo state, Nigeria

**DOI:** 10.1101/2025.02.27.25323070

**Authors:** Salako Tokunbo Abiodun, Akingbade Retta

## Abstract

A cross-sectional research design was utilized to investigate factors that influence organizational commitment. Participants comprised 397 healthcare professionals consisting of doctors, nurses, pharmacists, technicians, and administrative staff in public hospitals in Ibadan. Participants were selected using a multi-stage sampling technique. Data was collected through structured questionnaires. Participants’ ages ranged from 25 to 64 years, with a mean age of 41.58 years (SD = 9.90). Results showed that workplace happiness significantly influenced affective organizational commitment (*t* (395) = − 2.993, *p* < 0.01, *d* = −.30). Healthcare workers with high workplace happiness (x^-^ = 24.81, *SD* = 6.13) exhibited stronger affective commitment compared to those with low workplace happiness (*x*^-^ = 22.92, *S.D* = 6.44) Similarly, workplace happiness significantly influenced normative commitment (*t* (395) = − 2.603, *p* < 0.01, *d* = −.26), with employees who reported higher workplace happiness (*x*^-^ = 2.603, *p* < 0.01, *d* = −.26) showing greater normative commitment than those with lower workplace happiness (*x*^-^ = 24.40, *S D* = 5.46) However, there was no significant influence of workplace happiness on the continuance dimension of organisational commitment (*t* (395) = − 1.898, *p* =.058). Findings suggest workplace happiness is crucial for fostering affective and normative commitment. Healthcare organizations should enhance workplace happiness through supportive environments, recognition, and open communication to strengthen affective and normative commitment.

## Introductions

Organisational commitment among healthcare workers is crucial for effective healthcare delivery and positive patient outcomes (Al Otaibi, et al., 2023). While high-pressure environments are common across various sectors, the healthcare sector faces unique challenges where worker commitment directly impacts system functionality and quality of care. Healthcare professionals, including doctors, nurses, pharmacists, and support staff, operate in inherently stressful and emotionally demanding settings (Aruoture & Adegbie, 2024)). These pressures are compounded by external factors such as resource limitations and ongoing global health crises, as exemplified by the COVID-19 pandemic. Organizational commitment, defined as the emotional and psychological attachment to one’s workplace, comprises affective, continuance, and normative dimensions (Meyer & Allen, 1991). Affective commitment, the emotional connection and identification with the organization, is often driven by meaningful work aligned with personal values (Al Otaibi, et al., 2023). In collectivist cultures like Nigeria, healthcare workers frequently view their roles as serving their communities, fostering a sense of purpose (Obi, 2020). However, inadequate infrastructure, delayed salaries, and poor recognition can lead to emotional detachment and reduced commitment (Okon & Ede, 2021).

Continuance commitment, the recognition of costs associated with leaving the organization, can be influenced by economic realities and limited job opportunities. In Nigeria, this may be particularly relevant due to the global shortage of healthcare workers and the challenges of international relocation (Abelsen, et al., 2020). While continuance commitment may ensure retention, it does not necessarily equate to job satisfaction. For example, healthcare workers in rural areas may remain due to a lack of alternatives, but without a supportive work environment, they may experience low morale and burnout, negatively impacting service delivery (Adebola et al., 2019). Normative commitment, based on a sense of obligation, can be strong in the Nigerian healthcare system due to cultural expectations, professional ethics, and loyalty to the community (Nwankwo, et al., 2022). Many healthcare workers feel a moral duty to provide care, especially in underserved areas. Despite challenging conditions like resource constraints, infectious disease exposure, and security risks, this sense of obligation motivates many to remain in their roles. However, relying solely on normative commitment without addressing systemic issues can lead to resentment and diminished long-term commitment (Udenigwe, et al., 2022).

Organizational commitment is paramount in healthcare due to the critical nature of its operations. Lack of commitment can lead to substandard patient care, increased medical errors, and poorer health outcomes. Research demonstrates a strong link between happiness, job satisfaction, commitment, and performance (Ebeye et al., 2013). Positive emotional states among healthcare workers improve organizational outcomes through enhanced resilience and collaboration. Despite this, many healthcare professionals report dissatisfaction and disconnection due to structural and psychosocial factors. The Nigerian healthcare sector faces numerous challenges, including government underfunding, healthcare worker dissatisfaction, and brain drain. Inadequate budgetary allocations, delayed salaries, and insufficient infrastructure are persistent issues (Aruoture & Adegbie, 2024; Akinwale & George, 2023). Insufficient remuneration, suboptimal working conditions, and limited access to technology drive migration to better-resourced foreign systems, contributing to over one billion dollars spent annually on medical tourism. While collectivist values foster a sense of duty, this intrinsic motivation is often insufficient to overcome systemic challenges. This study examines the influence of workplace happiness on organizational commitment within this context.

Workplace happiness, encompassing positive emotional states, satisfaction, engagement, and well-being, is crucial for employee performance and organizational success (Kun & Gadanecz, 2022; Javanmardnejad, et al., 2021). It involves a proactive sense of contentment and motivation, driven by factors like fair compensation, growth opportunities, supportive leadership, work-life balance, and recognition (Akinwale, et al., 2024; Ekpechi & Igwe, 2023). This is particularly important in the healthcare sector, where professionals experience higher rates of distress, burnout, depressive symptoms, and suicidal ideation compared to other sectors. Employers strive to cultivate workplace happiness to enhance employee relations, improve remuneration, and retain competent staff, ultimately boosting organizational performance. Workplace happiness is paramount among workers in the healthcare sector. For instance, there is more evidence that healthcare professionals experience higher levels of distress, burnout syndrome, depressive symptoms, and suicidal ideations than other professionals in other sectors. Employers attempt to keep employees happy in order to enhance good relationships among employees and employment remunerations and to retain competent and productive (; Ekpechi & Igwe, 2023). The happiness of employees has a positive impact on their mindset; perform optimally which helps employers to achieve sales, production and sales targets (Javanmardnejad, et al., 2021).

Several interconnected factors contribute to low organizational commitment among healthcare workers in Nigeria. Economic issues, such as inflation and inadequate remuneration, foster feelings of unfairness and stress (Onah et al., 2022). Social comparisons with better-compensated colleagues in other countries exacerbate dissatisfaction and fuel brain drain (Ballard et al., 2021). Resource constraints, infrastructural decay, and limited support systems further compound these challenges. Fairness is often lacking due to inequitable policies regarding resource distribution, promotions, and compensation, undermining commitment and productivity (Faramarzpour et al., 2021; Rasheed et al., 2020; Chidi et al., 2023). Government underfunding, exemplified by the consistently low health budget allocation (Adebisi et al., 2020; Owoye & Onafowora, 2023), creates an environment where healthcare workers feel undervalued and unsupported.

The persistent insecurity in Nigeria, particularly the rise in kidnappings, significantly impacts various sectors, notably healthcare (Akinyemi et al., 2022). Healthcare workers, vital to national well-being, face threats, abductions, and unsafe working conditions, exacerbating existing healthcare system challenges. These threats contribute to a precarious environment that undermines organizational commitment, a crucial element for effective healthcare institutions. Low organizational commitment among Nigerian healthcare workers is a pressing concern with significant implications for service delivery and patient care. Burnout and dissatisfaction are prevalent among healthcare workers due to systemic inefficiencies, resource shortages, and overwhelming workloads (Orunbon et al., 2022). These challenges lead to frustration and detachment, driving many to seek opportunities abroad or in other industries, fueling brain drain. This exodus worsens workforce shortages and diminishes the morale of remaining employees, creating a cycle of low commitment and high turnover. Organizational commitment, encompassing employee loyalty, emotional attachment, and identification with their workplace, is essential for maintaining quality healthcare. The inability to retain a committed workforce jeopardizes healthcare institution efficiency (Ahmed et al., 2021; Anwar & Abdullah, 2021).

These challenges are compounded by poor working conditions, inadequate remuneration, limited career growth, and insufficient resources (Akinwale & George, 2023; Bolan et al., 2021; Nwankwo et al., 2021). These factors collectively undermine employee loyalty and dedication. Despite the recognized importance of organizational commitment, targeted strategies to address this issue remain limited. While existing literature highlights physical challenges, there is insufficient exploration of the psychological and emotional dimensions influencing commitment, particularly in the Nigerian context. Studies have shown that positive psychological environments promote emotional attachment, loyalty, and reduced turnover (Naz et al., 2020; Ogunbanjo et al., 2022). However, these studies often focus on academic staff, leaving a gap in understanding these dynamics within healthcare. Despite these insights, limited attention has been given to how workplace happiness affects organizational commitment among Nigerian healthcare workers. This study aimed to address these gaps by investigating the influence of workplace happiness on organizational commitment among healthcare workers in Nigeria.

The objective of this study was to determine if workplace happiness will influence organizational commitment among healthcare workers in the Ibadan metropolis. From the literature review, the following hypotheses were tested

1. Participants who report higher Workplace Happiness will express higher Affective Organisational Commitment than those who report lower Workplace Happiness.
2. Participants who report higher Workplace Happiness will express higher Continuance Organisational Commitment than those who report lower Workplace Happiness.
3. Participants who report higher Workplace Happiness will express higher Normative Organisational Commitment than those who report lower Workplace Happiness.
4. Participants who report higher Workplace Happiness will express higher Organisational Commitment than those who report lower Workplace Happiness.

## Method

### Participants and Procedure

Participants for this study were clinical and administrative staff employed in public hospitals in Ibadan, Oyo State, Nigeria. Three hundred and ninety-seven healthcare workers were selected from a population of 3,496 public health workers (Human Resource Office, 2022). The sample size was calculated using the Taro Yamane formula (Yamane, 1973). The selection of public hospitals was based on the peculiar challenges faced by such institutions, such as limited resources and staffing shortages, which can impact organizational commitment. Oyo State was chosen for its well-established healthcare system, offering a diverse and structured sample. Participants’ ages ranged from 25 to 64 years, with a mean age of 41.58 years (SD = 9.90).

To ensure a representative sample of healthcare professionals across various specializations, multi-stage sampling technique was employed. In the first stage, purposive selection was made of five government-owned hospitals in Ibadan, Oyo State. The hospitals were selected due to their prominent role in healthcare provision within the state. In the second stage, stratified random sampling was used to select participants from both clinical and administrative staff within each hospital. Stratification ensured representation from various professional groups, including doctors, nurses, pharmacists, and allied health professionals. Random sampling within each stratum was applied to maintain a representative sample.

A total of 435 questionnaires, of which 397 were adequately filled and analyzed. Prior to data collection, the researcher sought approval from the relevant departments in the selected hospitals. This approval facilitated distribution and retrieval of the questionnaires. Participants were provided with an informed consent form, which included a summary of the study and their rights as participants. Those who consented were administered the questionnaire. Participants were assured of anonymity and confidentiality of responses. They were also informed that they could withdraw from the study at any time. A written consent form was signed by the participants, acknowledging their understanding and agreement to participate.

### Measures

The instrument for this study was a structured questionnaire with four sections. Section one gathered demographic information. Section Two measured Organizational Commitment, Section Three measured Workplace Happiness. Organizational commitment was assessed using the Organizational Commitment (OC) Scale developed by Meyer and Allen (1997). This scale measures organizational commitment across three dimensions: affective commitment, which reflects employees’ emotional attachment to their organization; continuance commitment, which assesses the perceived costs associated with leaving the organization; and normative commitment, which captures employees’ feelings of obligation to remain with the organization. The OC Scale consists of 18 items, with six items assigned to each dimension, and responses are rated on a 7-point Likert scale. Cronbach’s alpha reliability coefficients for the scale ranges from 0.77 to 0.89 across different studies, indicating high internal consistency (Meyer & Allen, 1997).

Workplace happiness was measured using the Shortened Happiness at Work (SHAW) Scale, which evaluates employee engagement and work satisfaction. The SHAW scale comprises six items rated on a 5-point Likert scale, ranging from strongly disagree (1) to strongly agree (5), with higher scores reflecting greater happiness at work. It provides a concise yet reliable measure of workplace happiness, the scale has demonstrated strong psychometric properties, with reported reliability coefficients exceeding 0.80 in previous studies, supporting its internal consistency and validity. Data was analyzed using the Independent Sample’ t-test.

## Results

Three hundred and ninety-seven (397) respondents participated in the study. Gender comprised of 176 males (44.3%) and 219 females (55.2%); other gender was 14 (0.5%). Ages of participants ranged from 25 to 58 years (Mean = 41.58, SD = 8.90). Length of service shows the years spent on the job ranged from 2 to 37 years (Mean = 15.01, SD = 8.42). Approximately 25 (6.3%) of the respondents work in Adeoyo Memorial Specialist Hospital, 31 (7.8%) work in Adeoyo Yemetu Hospital, 22 (5.5%) work in Jericho Nursing Home, 36 (9.1%) work in Jericho Specialist Hospital and 283 (71.3%) work in University College Hospital, Ibadan. Marital status of participants showed that 12.1% (48) the respondents were single, 83.1% (330) were married, and 4.8% (19) were divorced. Approximately 37.3% (148) were Doctors, 10.3% (41)were pharmacists, 26.2% (104) were Nurses, 6.3% (25) were Technicians and 19.9% (79) were Administrators.

The results in Table 1 show that there is a significant difference between low workplace happiness 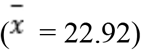 and high workplace happiness 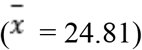 and affective organizational commitment (t (395) = -2.993, p < 0.01). This means healthcare workers who have high workplace happiness exhibited higher affective organizational commitment than healthcare workers who have low workplace happiness. The effect size was calculated with Cohen’s d, and shows that the effect size is moderate (d = -.30).

**Table 1.** Summary of t-test for independent sample showing the difference between low and high work happiness on and affective organizational commitment.

| Variable | N | $\bar{x}$ | SD | df | t | p | d |
| --- | --- | --- | --- | --- | --- | --- | --- |
| Low workplace happiness | 195 | 22.92 | 6.44 | 395 | -2.993 | < .01 | -.30 |
| High workplace happiness | 202 | 24.81 | 6.13 |  |  |  |  |

The results in Table 2 show that there is no significant difference between low workplace happiness 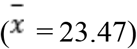 and high workplace happiness 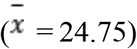 on continuance organizational commitment (t (395) = -1.898, p > 0.05). This means healthcare workers who have high workplace happiness did not exhibit higher affective organizational commitment than healthcare workers who have low workplace happiness.

**Table 2.**
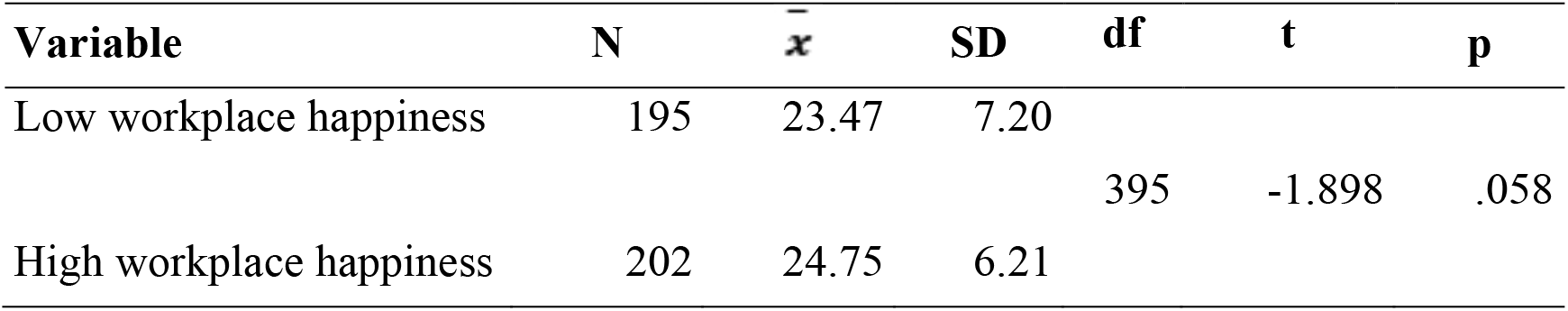
Summary of t-test for independent sample showing the difference between low and high work happiness and continuance organizational commitment.

The results in Table 3 show that there was a significant difference between low workplace happiness 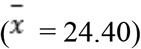 and high workplace happiness 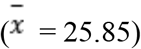 on normative organizational commitment (t (395) = -2.603, p < 0.01). This means healthcare workers who have high workplace happiness exhibited higher normative organizational commitment than healthcare workers who have low workplace happiness. The effect size was calculated with Cohen’s d, and this shows that the size of effect of workplace happiness on normative organizational commitment is moderate (d = -.26).

**Table 3:**
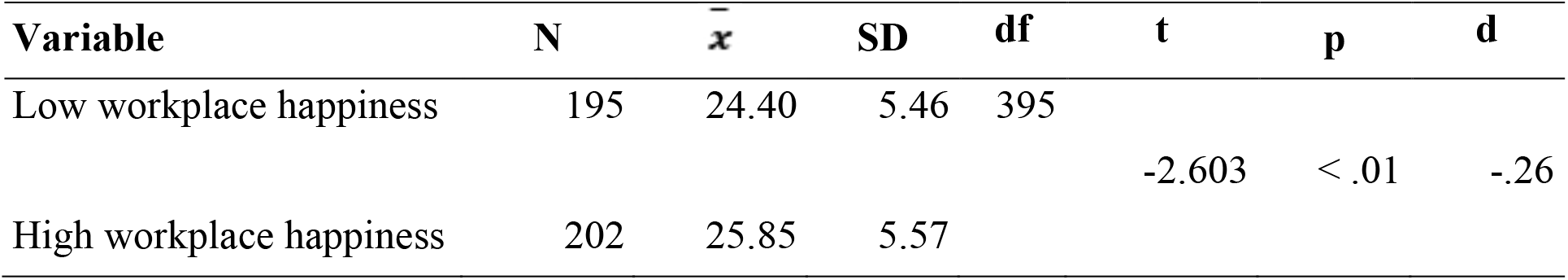
Summary of t-test for independent sample showing the difference between low and high work happiness and normative organizational commitment.

| Variable | N | $\bar{x}$ | SD | df | t | p | d |
| --- | --- | --- | --- | --- | --- | --- | --- |
| Low workplace happiness | 195 | 24.40 | 5.46 | 395 | -2.603 | < .01 | -.26 |
| High workplace happiness | 202 | 25.85 | 5.57 |  |  |  |  |

The results in Table 4 show that there is no significant difference between low workplace happiness 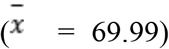 and high workplace happiness 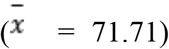 on organizational commitment (t (395) = -1.305, p > 0.05). This means healthcare workers who have high workplace happiness did not exhibit higher organizational commitment than healthcare workers who have low workplace happiness.

**Table 4:** Summary of t-test for independent sample showing the difference between low and high work happiness on organizational commitment.

| Variable | N | $\bar{x}$ | SD | df | t | p |
| --- | --- | --- | --- | --- | --- | --- |
| Low workplace happiness | 195 | 69.99 | 14.36 | 395 | -1.305 | > .05 |
| High workplace happiness | 202 | 71.71 | 11.95 |  |  |  |

## Discussion

This study investigated the influence of workplace happiness on organisational commitment among healthcare workers. The following hypotheses were tested

i. Participants who report higher Workplace Happiness will express higher Affective Organisational Commitment than those who report lower Workplace Happiness.
ii. Participants who report higher Workplace Happiness will express higher Continuance Organisational Commitment than those who report lower Workplace Happiness.
iii. Participants who report higher Workplace Happiness will express higher Normative Organisational Commitment than those who report lower Workplace Happiness.
iv. Participants who report higher Workplace Happiness will express higher Organisational Commitment than those who report lower Workplace Happiness.

The Four hypotheses were tested and the findings are as follows:

The findings of the first hypothesis indeed revealed a significant difference in affective organizational commitment of healthcare workers. Individuals with higher levels of workplace happiness demonstrated greater affective organizational commitment. This result implies that workplace happiness plays an important role in enhancing employees’ emotional attachment and identification with their organization. Healthcare workers who experience greater happiness in their work environment are more likely to feel committed to their organization, reflecting a deeper sense of loyalty and motivation to contribute positively.

Previous research has demonstrated a relationship between positive work environments and increased employee loyalty. Studies by Akgunduz et al. (2023) and Kustiawan et al. (2022) highlight the significance of workplace happiness, encompassing job satisfaction, engagement, and a sense of purpose, in fostering employee commitment. Happy and fulfilled employees are more likely to develop a strong emotional connection to their organization, leading to increased affective commitment. However, Ahmad et al. (2024) emphasize the influence of emotional experiences on affective commitment. Positive emotions, often arising from workplace happiness, contribute to a sense of belonging and a desire to remain with the organization (Al Halbusi et al., 2023). This aligns with the current findings, where healthcare workers experiencing higher levels of workplace happiness demonstrated greater emotional commitment to their organizations. Further evidence is provided by Babatunde and Magret (2023), who argue that workplace happiness is often a result of job resources such as support, autonomy, and professional growth opportunities. These resources not only enhance employee well-being but also strengthen their organizational commitment by creating supportive and fulfilling work environments.

The second hypothesis which proposed that participants with higher workplace happiness would demonstrate a higher continuance organisational commitment compared to those with lower workplace happiness. However, the results indicate no significant difference between healthcare workers who reported high workplace happiness and those who reported low workplace happiness in terms of their continuance organizational commitment. This suggests that workplace happiness may not play a critical role in influencing this dimension of organizational commitment. Continuance organizational commitment is typically characterized by an employee’s perceived need to remain with an organization due to the costs associated with leaving, such as loss of financial benefits, job security, or limited alternative opportunities. Unlike affective commitment, which is driven by emotional attachment, continuance commitment is more calculative and pragmatic. The findings imply that the emotional benefits derived from workplace happiness do not strongly impact the decision to stay in an organization based on these calculative factors.

This study is supported by the findings of Ardo et al. (2024) and Chiedu et al. (2024), who discovered that while characteristics like as job satisfaction and workplace happiness were strong predictors of affective commitment, their effect on continuation commitment was limited. Instead, continued commitment was more significantly associated with career alternatives, longevity, and individuals’ perceived investments in their employment. Islam et al., (2023) discovered that continuous commitment is mostly driven by calculative considerations, such as a lack of other career prospects and fear of financial loss, rather than workplace satisfaction or good feelings. Kustiawan et al. (2022) reiterated that employees’ decision to stay or leave an organization often depends on a cost-benefit analysis. While workplace factors like happiness and job satisfaction can influence emotional attachment (affective commitment), they have a weaker impact on the decision to stay based on economic considerations (continuance commitment).

The third hypothesis states that healthcare workers with higher workplace happiness will exhibit higher normative organizational commitment compared to those with lower workplace happiness. The results confirm this hypothesis, revealing a significant difference between the two groups. Healthcare workers who report high levels of workplace happiness demonstrate a greater sense of normative organizational commitment than their counterparts with lower workplace happiness. This indicates that workplace happiness positively influences the sense of moral obligation or duty to remain with the organization. Empirical evidence supports the idea that workplace happiness significantly influences employees’ organizational commitment. Research by Gumasing, and Ilo, (2023) and Kaushal, (2020) describes job satisfaction and happiness at work as a positive emotional state derived from workplace experiences. This satisfaction serves as a precursor to various forms of organizational commitment, including normative commitment. Employees who are more satisfied with their jobs tend to be more engaged, leading to a stronger commitment to organizational goals and values (Al-Refaei et al., 2023). Other studies by Taştan et al. (2020) and Thompson and Bruk-Lee (2021) support this link, demonstrating that workplace happiness is a predictor of organizational commitment.

The fourth hypothesis posited that participants with higher workplace happiness would demonstrate greater organizational commitment compared to those with lower workplace happiness. However, the analysis did not yield statistically significant results, indicating that healthcare workers who reported higher workplace happiness did not exhibit substantially higher organizational commitment than their less happy counterparts. This result implies that while workplace happiness might contribute to various aspects of employee experience, it does not necessarily translate into a universal increase in overall organizational commitment. To support this finding, Oyelakin et al., (2021) and George, (2021) discovered a significant positive effect of workplace happiness on organisational commitment, implying that when employees feel happy and fulfilled at work, they are more likely to develop a strong sense of loyalty and responsibility to their organisation. Furthermore, Babatunde and Magret (2023) demonstrated a link between higher levels of happiness and increased normative commitment among employees. Employees who reported higher job satisfaction and overall happiness were more likely to express loyalty to their organization. Akgunduz et al. (2023) showed that happier employees are less likely to leave their organizations and more likely to engage in behaviours beneficial to the organization, such as helping colleagues and promoting the organization.

## Conclusion

The findings of this study highlight the influence of workplace happiness on organizational commitment among healthcare workers in Ibadan. Specifically, the results indicate that workplace happiness has a significant influence on certain dimensions of organizational commitment, particularly affective and normative commitment. Healthcare workers who reported higher levels of workplace happiness exhibited a stronger emotional attachment to their organization, suggesting that a positive and supportive work environment fosters deeper organizational loyalty. Similarly, the study found that workplace happiness contributed to a greater sense of obligation or normative commitment, implying that when employees feel valued and satisfied at work, they are more likely to develop a sense of duty toward their organization. However, the results also reveal that workplace happiness did not have a significant impact on continuance commitment, which is primarily driven by practical considerations such as financial security and job stability. This suggests that while workplace happiness enhances emotional and moral attachment to the organization, it may not necessarily influence employees’ decisions to remain in their jobs based on necessity or perceived costs of leaving. Furthermore, the overall measure of organizational commitment did not show a significant difference between employees with low and high workplace happiness, indicating that other factors beyond workplace happiness, such as job security, professional growth opportunities, and workplace policies, may play a more dominant role in determining long-term commitment.

## Implications of the Study

The findings of this study have significant implications for healthcare management, employee engagement strategies, and organizational policies. The study highlights that workplace happiness plays a crucial role in strengthening affective and normative commitment among healthcare workers. This suggests that when employees experience a positive work environment, they are more likely to develop an emotional bond with their organization and feel a sense of duty to remain committed. However, since workplace happiness did not significantly influence continuance commitment, it implies that factors such as job security, financial benefits, and career advancement opportunities may play a greater role in determining whether healthcare workers stay in their jobs out of necessity. This indicates that while fostering workplace happiness is important, it must be complemented with structural and policy-driven efforts that address employees’ long-term career stability. Additionally, the findings suggest that healthcare organizations should prioritize employee well-being to enhance overall job satisfaction and organizational loyalty. Since the healthcare sector is often characterized by high levels of stress, long working hours, and emotional exhaustion, promoting workplace happiness can help mitigate burnout and improve job performance. The study also has broader implications for employee retention strategies, as it suggests that focusing on workplace happiness alone may not be enough to prevent turnover. Instead, a holistic approach that integrates both emotional well-being and financial security is essential for strengthening organizational commitment in the healthcare sector.

## Recommendations

Based on these findings, several recommendations can be made to improve workplace happiness and organizational commitment among healthcare workers. First, healthcare organizations should implement policies that promote a positive work culture by fostering supportive leadership, recognizing employee contributions, and ensuring open communication channels. Creating an environment where employees feel valued and appreciated can enhance their emotional connection to the organization and encourage a stronger sense of belonging. Second, healthcare institutions should invest in employee well-being programs that address both mental and physical health needs. Providing regular access to counselling services, stress management workshops, and wellness initiatives can improve workplace happiness and reduce the likelihood of burnout. Additionally, incorporating flexible work schedules and ensuring manageable workloads can contribute to higher job satisfaction and increased affective commitment.

Furthermore, while workplace happiness is essential, organizations must also focus on job security and career development opportunities. Offering competitive salaries, professional growth programs, and career advancement pathways can help reinforce continuance commitment by reducing employees’ concerns about financial stability. Providing scholarships, training, and mentorship programs can also empower healthcare workers to see long-term prospects within their organizations. Also, healthcare institutions should develop tailored retention strategies that align with employees’ motivations and expectations. Conducting regular employee satisfaction surveys can provide insights into factors influencing workplace happiness and organizational commitment, enabling management to make data-driven decisions. By integrating both psychological well-being and structural support systems, healthcare organizations can create a more committed and engaged workforce, ultimately improving service delivery and patient care outcomes.

## Limitations and Suggestions for further studies

This study has several limitations that should be acknowledged. First, the cross-sectional design limits the ability to establish causal relationships between workplace happiness and organizational commitment. Longitudinal studies would provide a clearer understanding of how these factors influence commitment over time. Second, the study relied on self-reported data, which may be subject to social desirability bias, potentially affecting the accuracy of responses. Future research could incorporate qualitative methods, such as in-depth interviews, to gain richer insights into the experiences of healthcare workers. Additionally, the study focused on healthcare workers in a specific region, which may limit the generalizability of the findings to other contexts, particularly in different healthcare settings or countries with varying workplace cultures. Further studies should explore how workplace happiness interacts with other factors, such as leadership styles, job satisfaction, and employee retention, to provide a more comprehensive understanding of organizational commitment in the healthcare sector.

## Data Availability

All data produced in the present study are available upon reasonable request to the authors.

